# Kleine Levin syndrome is associated with birth difficulties and genetic variants in the TRANK1 gene loci

**DOI:** 10.1101/2021.01.08.20249006

**Authors:** Aditya Ambati, Ryan Hillary, Smaranda Leu-Semenescu, Hanna M. Ollila, Ling Lin, Emmanuel During, Neal Farber, Thomas J Rico, Juliette Faraco, Eileen Leary, Andrea Goldstein-Piekarski, Yu-Shu Huang, Fang Han, Yakov Sivan, Michel Lecendreux, Pauline Dodet, Makoto Honda, Natan Gadoth, Sona Nevsimalova, Fabio Pizza, Takashi Kanbayashi, Rosa Peraita Adrados, Guy Leschziner, Rosa Hasan, Francesca Canellas, Kazuhiko Kume, Makrina Daniilidou, Patrice Bourgin, David Rye, José L Vicario, Birgit Högl, Seung Chul Hong, Guiseppe Plazzi, Geert Mayer, Anne Marie Landtblom, Yves Dauvilliers, Isabelle Arnulf, Emmanuel Mignot

**Affiliations:** Stanford University Center for Sleep Sciences and Medicine, Department of Psychiatry and Behavioral Sciences. 3165 Porter Drive, Palo Alto, CA 94304, USA; Sleep Disorders Unit (Department “R3S”), Pitié-Salpêtrière Hospital, APHP, National Reference Center for Narcolepsy, Idiopathic Hypersomnia and Kleine–Levin Syndrome, Sorbonne University, IHU@ICM, INSERM, CNRS UMR7225, MOV’IT team, F-75013 Paris, France; Institute for Molecular Medicine Finland, HiLIFE, University of Helsinki, Helsinki, Finland; Kleine-Levin Syndrome Foundation, Boston, MA, USA; Department of Child Psychiatry and Sleep Center, Chang Gung Memorial Hospital and University, Taoyuan, Taiwan; Department of Pulmonary Medicine, the Peking University People’s Hospital, Beijing, China; Safra Children’s Hospital, Sheba Medical Center, Tel Aviv University, Sackler Faculty of Medicine, Tel Aviv, Israel; Pediatric Sleep Center and National Reference Center for Narcolepsy and Idiopathic Hypersomnia Hospital Robert Debre, Paris, France; Tokyo Metropolitan Institute of Medical Science, Sleep Disorders project, Tokyo, Japan; Dept Neurology, Maynei Hayeshua Medical Center, Bnei Barak, and Sackler Faculty of Medicine, Tel-Aviv University, Israel; Department of Neurology, Charles University, 1st Faculty of Medicine and General Teaching Hospital, Prague, Czech Republic; Department of Biomedical and Neuromotor Sciences, University of Bologna and IRCCS Institute of Neurological Sciences, Bologna, Italy; Akita University, Department of Neuropsychiatry, Akita-City, Akita, 010-8543, Japan; Sleep and Epilepsy Unit – Clinical Neurophysiology Service, University General Hospital Gregorio Marañón, Research Institute Gregorio Marañón. University Complutense of Madrid, Spain; Sleep Disorders Centre, Guy’s Hospital, London SE1 9RT, England; Institute of Psychiatry, Hospital das Clinicas, Faculty of Medicine, University of Sao Paulo. Dr Ovidio Pires de Campos n. 785, 05403-010, Brazil; Department of Psychiatry, Hospital Universitari Son Espases, Palma de Mallorca, Health Research Institute of the Balearic Islands (IdISBa), Spain; Department of Neuropharmacology, Graduate School of Pharmaceutical Sciences, Nagoya, Japan; Division of Neurology, Department of Clinical and Experimental Medicine, University of Linköping, Linköping, Sweden/Department of Neuroscience/Neurology, Uppsala University, Uppsala, Sweden; Sleep disorders Center-CIRCSom, CHU Strasbourg & CNRS - Institute for Cellular and Integrative Neurosciences, University of Strasbourg; Emory Program in Sleep, Brain Health Center, Atlanta, GA USA; JLV Histocompatibility, Blood Center of the Community of Madrid, Madrid, Spain; Department of Neurology, Innsbruck Medical University, Austria; Department of Neuropsychiatry, St. Vincent’s Hospital, Catholic University of Korea, College of Medicine, Seoul, Korea; Hephata Klinik, Schimmelpfengstr. 6, 34613 Schwalmstadt, Germany; Philipps Universität Marburg, Baldinger Str., 35043 Marburg, Germany; Centre de référence nationale narcolepsie et hypersomnies rares, 34295 Montpellier cedex 5, France; Unité des troubles du sommeil et de l’éveil, hôpital Gui-De-Chauliac, 80, avenue Augustin-Fliche, 34295 Montpellier cedex 5, France; Inserm U1061, 34295 Montpellier, France

**Keywords:** Kleine-Levin Syndrome, KLS, Bipolar disorder, Schizophrenia, TRANK1, Birth Difficulties

## Abstract

Kleine-Levin Syndrome (KLS) is a rare disorder characterized by severe episodic hypersomnia, with cognitive impairment accompanied by apathy or disinhibition. Pathophysiology is unknown, although imaging studies indicate decreased activity in hypothalamic/thalamic areas during episodes. Familial occurrence is increased, and risk is associated with reports of a difficult birth. We conducted a worldwide case-control genome wide association study in 673 KLS cases collected over 14 years, and ethnically matched 15,341 control individuals. We found a strong genome-wide significant association (OR=1.48,rs71947865,p=8.6×10^−9^) with 20 single nucleotide polymorphisms encompassing a 35kb region located in the 3’ region of *TRANK1* gene, previously associated with bipolar disorder and schizophrenia. Strikingly, KLS cases with *TRANK1* rs71947865 variant had significantly increased reports of a difficult birth. As perinatal outcomes have dramatically improved over the last 40 years, we further stratified our sample by birth years and found that recent cases had a significantly reduced *TRANK1* rs71947865 association. While the*TRANK1* rs71947865 association did not replicate in the entire follow-up sample of 171 KLS cases, the TRANK1 rs71947865 was significantly associated with KLS in the subset follow-up sample of 59 KLS cases who reported birth difficulties (OR=1.54;p=0.01). Genetic liability of KLS as explained by polygenic risk scores was increased (pseudo r^2^=0.15;p<2.0×10^−22^ at p=0.5 threshold) in the follow-up sample. Pathway analysis of genetic associations identified enrichment of circadian regulation pathway genes in KLS cases. Our results suggest links between KLS, behavioral rhythmicity, and bipolar disorder, and indicates that the *TRANK1* polymorphisms in conjunction with reported birth difficulties may predispose to KLS.

**Significance Statement:** Genetic markers in *TRANK1* gene and its vicinity have been weakly associated with bipolar disorder and schizophrenia (10% increased risk). We found that the same polymorphisms are associated with Kleine-Levin Syndrome (50% increased risk), a rare sleep disorder characterized by recurrent episodes of severe hypersomnia and cognitive abnormalities. Response to lithium treatment are suggestive of a pathophysiological overlap between KLS and bipolar disorder. The study also shows that variants in the *TRANK1* gene region may predispose to KLS when patients have had a difficult birth, suggesting that *TRANK1* gene region modulate newborns’ response to brain injury, with consequences for mental and neurological health in adulthood. Another possibility may be that the polymorphism impact birth and KLS.

## Introduction

Kleine Levin Syndrome (KLS) was identified as a unique disease entity almost a century ago (1, 2). It affects primarily male teenagers and is characterized by unpredictable periods (median length ∼10 days) of intense hypersomnia (≥18 hours of sleep per day), occasionally associated with megaphagia and sexual disinhibition. More recent descriptions in large case series have confirmed a stereotypic appearance of abrupt onset episodes in mostly male (66-75%) patients and association of severe hypersomniac episodes with cognitive and behavioral disturbances (mental slowness, confusion, apathy, derealization, altered perception, and disinhibited behavior). Remarkably, there is a complete absence of the symptoms between episodes, and a generally favorable evolution, with spontaneous disappearance of episodes after 1-2 decades (3-6). KLS episodes are dramatic in appearance and patients are fully incapacitated. EEG studies occasionally reveal non-specific EEG slowing during episodes. Further, lithium has been shown to reduce episodes in about one-third of cases (4, 7). Episodes may be associated with flu-like symptoms at the onset, with seasonal winter occurrence suggested by some (8).

Although most cases of KLS are sporadic, familial occurrence (3-8% of relatives) has been reported in studies (5, 9) and there are reports of concordant twin pairs (9, 10)which, considering the rarity of the condition (a few cases per million individuals) (11), suggests genetic or shared environmental effects. Early studies had suggested an HLA association suggestive of an autoimmune mechanism (12), but this was not confirmed in larger samples (3, 8, 13). Further, karyotyping was normal in 112 patients, except for one patient with sporadic KLS who had a duplication in Xp22.31 (13).

This study is the first report of genetic associations in a discovery cohort of 673 KLS cases with attempted replication in 171 cases across the world. Additionally, targeted exome sequencing in 32 cases derived from 16 multiplex families (9) was conducted. A KLS Genome-Wide Association Study (GWAS) found a strong signal in the *TRANK1* region (rs71947865), similar to that reported in bipolar disorder and schizophrenia. In these latter disorders the TRANK1 association variably maps to rs9834970 in Caucasians or to rs3732386 in multi-ethnic populations, both SNPs located immediately 3’ of the TRANK1 gene (14-16). Strikingly, the *TRANK1* signal was mostly present in KLS patients reporting birth difficulties and decreased over time (defined by birth year), paralleling well-established improved perinatal care. Birth difficulties (and maternal infections) have also long been associated with increased risk of bipolar disorder and schizophrenia (17-29). These results indicate possible gene x environment interactions in KLS, with possibly similar effects in other TRANK1-associated disorders such as bipolar disorder and schizophrenia.

## Results

### Genome-Wide Association Studies (GWAS) of KLS

A worldwide multiethnic cohort of 673 KLS cases, the largest sample of KLS cases ever to be analyzed, was collected between the years of 2003 and 2017 and compared to 15,341 matched controls (the “discovery cohort”; see **Dataset S1** for sample construction, and **SI Appendix, fig. S1** for corresponding Principal Component Analysis-PCA plots). Genetic association analyses (genomic inflation λGC = 1.06) found a genome-wide significant effect (**Figure 1**) with no heterogeneity across ethnic groups, with more than 20 tightly linked single nucleotide polymorphisms (SNPs) encompassing a 35kb region of human chromosome 3 in the 3’ region of the tetratricopeptide repeat and ankyrin repeat-containing protein (*TRANK1*) gene. The TRANK1 gene was observed to have the highest expression in the fallopian tube, closely followed by the brain-cerebellum (GTEx v8) (see **Table 1, Figure 2** for zoom plot and **Dataset S2** where all variants in this locus are listed). The most significant association was observed at rs71947865 (OR=1.47, p=8.6×10^−9^, presence of G versus GGGAGCCA), although the highest OR was observed at rs6789885 (OR=1.53, p=8.9×10^−8^, T versus C, **Dataset S2**). This TRANK1 region SNP rs71947865 is in absolute Linkage Disequilibrium (LD) with rs3732386 (r^2^=1), a SNP associated with Bipolar Disorder and Schizophrenia in multiethnic samples (**Dataset S3**).

**Table 1:** Top variants in TRANK1 region associated with KLS in worldwide discovery cohort GWAS meta-analysis in 673 KLS cases and 15341 control individuals and in a follow-up replication cohort of 171 KLS cases and 1956 controls.Effect allele is B, genomic coordinates in hg19/GRCh37.

| Chr | SNP | BP | A/B | GWAS meta-analysis<br>n=673 |  |  | Follow-up samples<br>n=171 |  | Combined<br>n=844 |  |
| --- | --- | --- | --- | --- | --- | --- | --- | --- | --- | --- |
|  |  |  |  | Freq.B | OR | P value | OR | P value | OR | P value |
| 3 | rs71947865 | 36877798 | GGGAGCCA/G | 0.29 | 1.48 | 8.61x10<sup>-09</sup> | 1.17 | 0.19 | 1.40 | 3.51x10<sup>-8</sup> |
| 3 | rs3732385 | 36872193 | C/A | 0.30 | 1.47 | 1.44x10<sup>-8</sup> | 1.17 | 0.18 | 1.39 | 5.00x10<sup>-8</sup> |
| 3 | rs3732384 | 36872224 | T/C | 0.30 | 1.47 | 1.44x10<sup>-8</sup> | 1.17 | 0.18 | 1.39 | 5.00x10<sup>-8</sup> |
| 3 | rs7652637 | 36871507 | T/C | 0.30 | 1.47 | 1.47x10<sup>-8</sup> | 1.18 | 0.18 | 1.39 | 5.06x10<sup>-8</sup> |
| 3 | rs9821223 | 36883369 | T/C | 0.29 | 1.47 | 1.5x10<sup>-8</sup> | 1.11 | 0.39 | 1.38 | 1.67x10<sup>-7</sup> |
| 3 | rs9811916 | 36878086 | A/G | 0.30 | 1.47 | 1.63x10<sup>-8</sup> | 1.17 | 0.19 | 1.39 | 5.78x10<sup>-8</sup> |
| 3 | rs4678909 | 36878563 | A/G | 0.30 | 1.47 | 1.73x10<sup>-8</sup> | 1.16 | 0.21 | 1.39 | 7.42x10<sup>-8</sup> |
| 3 | rs6771655 | 36884370 | C/A | 0.29 | 1.47 | 1.74x10<sup>-8</sup> | 1.11 | 0.39 | 1.38 | 1.91x10<sup>-7</sup> |
| 3 | rs9882911 | 36885392 | T/C | 0.29 | 1.47 | 1.78x10<sup>-8</sup> | 1.11 | 0.39 | 1.38 | 1.96x10<sup>-7</sup> |
| 3 | rs9863798 | 36885960 | C/T | 0.29 | 1.47 | 1.99x10<sup>-8</sup> | 1.11 | 0.39 | 1.37 | 2.16x10<sup>-7</sup> |
| 3 | rs4678910 | 36887973 | C/G | 0.29 | 1.47 | 2.03x10<sup>-8</sup> | 1.11 | 0.41 | 1.37 | 2.38x10<sup>-7</sup> |
| 3 | rs4678911 | 36888064 | G/T | 0.29 | 1.47 | 2.09x10<sup>-8</sup> | 1.11 | 0.40 | 1.37 | 2.29x10<sup>-7</sup> |
| 3 | rs9819304 | 36888172 | T/G | 0.29 | 1.47 | 2.1x10<sup>-8</sup> | 1.11 | 0.40 | 1.37 | 2.30x10<sup>-7</sup> |
| 3 | rs17807744 | 36894048 | C/T | 0.29 | 1.46 | 2.18x10<sup>-8</sup> | 1.10 | 0.45 | 1.37 | 2.92x10<sup>-7</sup> |
| 3 | rs4234258 | 36878970 | A/G | 0.30 | 1.46 | 2.26x10<sup>-8</sup> | 1.16 | 0.23 | 1.38 | 9.95x10<sup>-8</sup> |
| 3 | rs3732386 | 36871993 | C/T | 0.30 | 1.46 | 2.26x10<sup>-8</sup> | 1.17 | 0.18 | 1.39 | 7.16x10<sup>-8</sup> |
| 3 | rs6550435 | 36864489 | T/G | 0.29 | 1.47 | 2.37x10<sup>-8</sup> | 1.17 | 0.20 | 1.39 | 8.62x10<sup>-8</sup> |
| 3 | rs17195098 | 36893965 | C/T | 0.29 | 1.46 | 2.83x10<sup>-8</sup> | 1.10 | 0.45 | 1.36 | 3.63x10<sup>-7</sup> |
| 3 | rs9883001 | 36894579 | C/T | 0.29 | 1.46 | 2.9x10<sup>-8</sup> | 1.10 | 0.45 | 1.36 | 3.70x10<sup>-7</sup> |
| 3 | rs4678912 | 36894646 | A/T | 0.29 | 1.46 | 2.91x10<sup>-8</sup> | 1.10 | 0.45 | 1.36 | 3.71x10<sup>-7</sup> |
| 3 | rs1532965 | 36861196 | T/G | 0.30 | 1.46 | 2.92x10<sup>-8</sup> | 1.16 | 0.22 | 1.38 | 1.20x10<sup>-7</sup> |
| 3 | rs4678913 | 36895741 | C/G | 0.29 | 1.46 | 3.05x10<sup>-8</sup> | 1.10 | 0.45 | 1.36 | 3.93x10<sup>-7</sup> |
| 3 | rs11706780 | 36896235 | C/T | 0.29 | 1.46 | 3.12x10<sup>-8</sup> | 1.09 | 0.46 | 1.36 | 4.04x10<sup>-7</sup> |
| 3 | rs4624519 | 36862980 | C/T | 0.30 | 1.45 | 3.39x10<sup>-8</sup> | 1.16 | 0.22 | 1.38 | 1.36x10<sup>-7</sup> |

**Figure 1:**
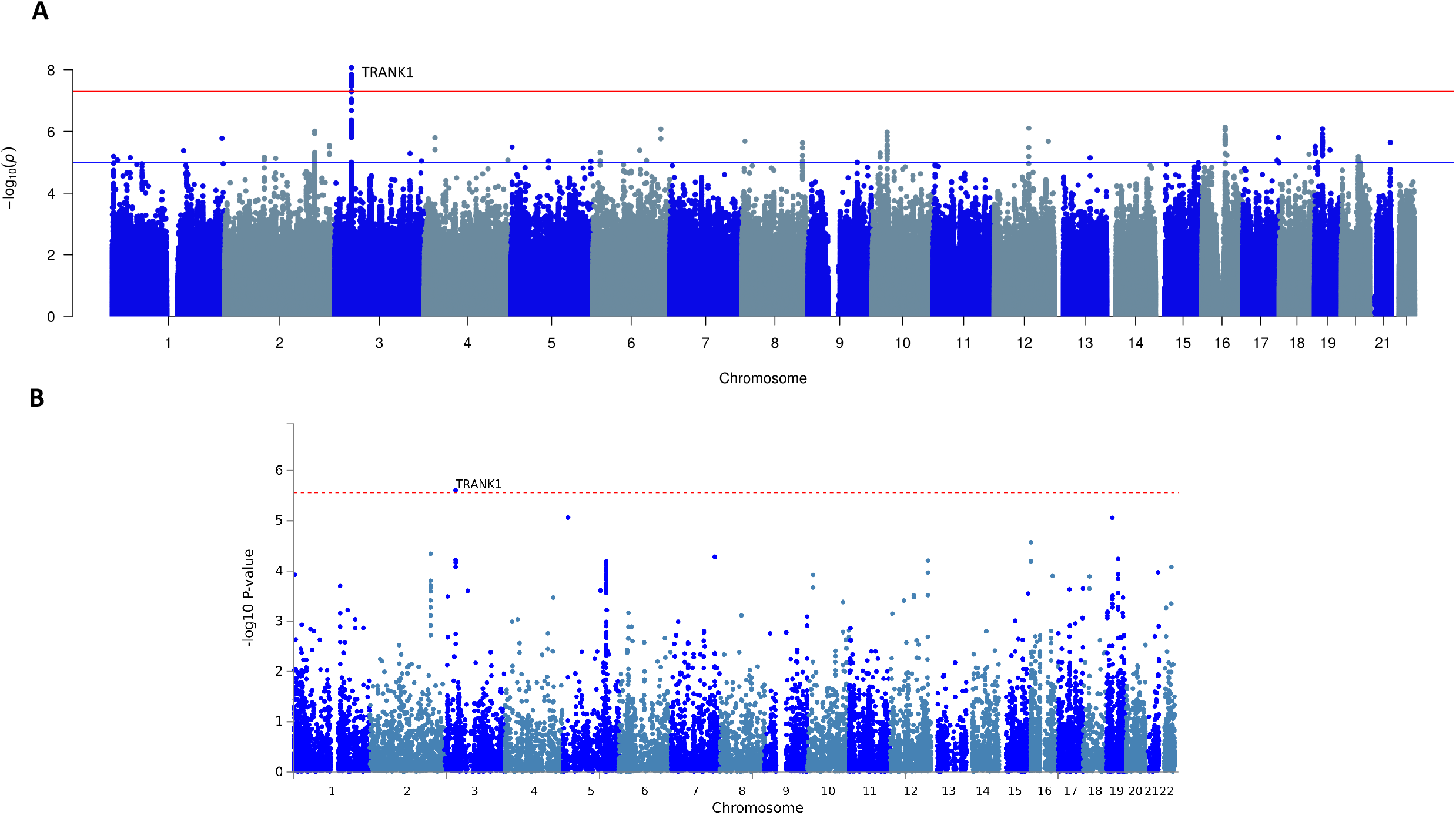
Multiethnic GWAS in 673 KLS cases and 15431 control individuals (A) Manhattan plot of the meta-analysis association results from SNP based GWAS study. The X-axis shows genomic location by chromosome and the y-axis shows -log10 p-values. Red horizontal line indicates genome-wide significant p-value threshold of 5×10^−8^. Individual loci are annotated. (B) Manhattan plot of gene-based association test association results, SNPs were mapped to 18247 protein coding genes (distance 0). Genome wide significance (red dashed line in the plot) was defined at P = 0.05/18247 = 2.74×10^−6^.

**Figure 2:**
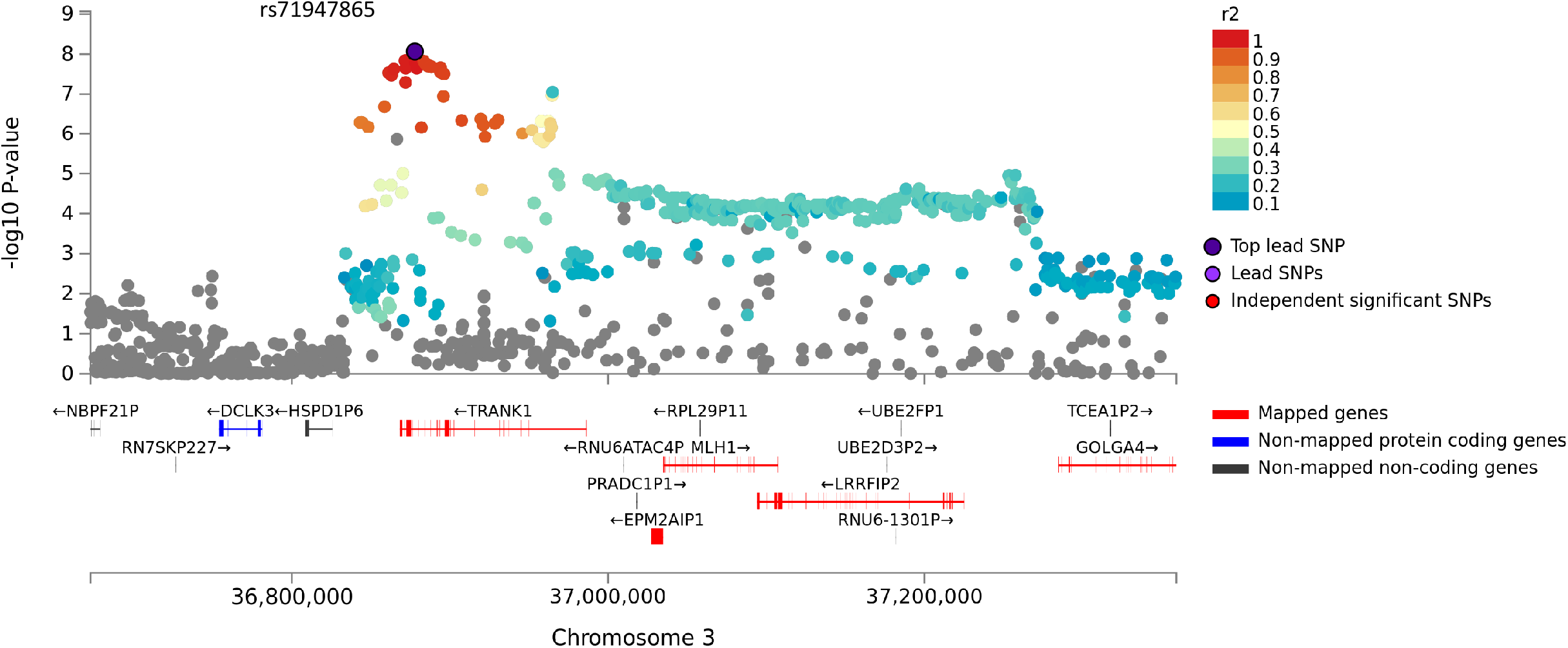
Regional association plot of the GWA significance of chromosome 3 flanked by rs76652637 and rs11706780 in the 3’’ region of *TRANK1*. Highest significance was observed at rs71947865 (OR=1.47, p=8.6×10^−9^, presence of G versus GGGAGCCA). The variants are color coded by the magnitude of LD with the rs71947865.

The Linkage Disequilibrium (LD) block contains several eQTL (expression quantitative trait loci) effects affecting numerous genes such as TRANK1, LRRFIP2, MLH1, GOLGA4, DCLK3, STAC and long non-coding RNA RP11-640L9.2. The SNP rs11129735 (KLS discovery GWAS p= 9.9×10^−6^; LD r^2^=0.4 with our top GWAS significant variants) had the highest coloc (CLPP) probability (>0.95 causal probability) using eCAVIAR, with the expression of LRRFIP2 in the thyroid, DCLK3 in skin and brain frontal cortex, C3orf35 in testis, MLH1 in breast tissue, TRANK1 in the spinal cord and EPM2AIP1 in the hypothalamus (GTEx v7). Querying the GTEx v8 database (30), we found our lead SNP rs71947865 variant to be a strong eQTL for LRRFIP2 gene expression in the thyroid (p=8.2×10^−20^) and TRANK1 expression in breast-mammary tissue and whole blood. rs71947865 also had multiple effects on LRRFIP2 expression in various tissues (p<10^−6^, GTEx portal v8). LRRFIP2 together with MLH1 and GOLGA4 are highly expressed in placenta and differentially expressed in placentae from complicated compared to normal pregnancies (31, 32). The tissue expression for most of these genes is widespread in the brain and poorly characterized. For the reason of convenience, we further called this effect “TRANK1-region polymorphisms” or “TRANK1 effect”, although data to date does not allow genetic attribution of the observed effects to secondary changes in TRANK1 gene expression.

### Association of symptoms with polymorphisms

Clinical data collected from 592 of 673 KLS patients from the discovery cohort were next scrutinized for genotype effects. Our *a priori* hypothesis for testing was to explore whether TRANK1 polymorphisms were associated with either more depressive symptoms, considering lithium responsiveness in the syndrome, and/or a differential effect on birth difficulties, a robust finding in our prior study (3). Other variables were compared as exploratory analyses. *TRANK1* rs71947865 (G) genotype was most significantly associated with reports of birth difficulties (OR=1.65; p<0.005 Fisher’s exact test of allelic counts) and developmental delays. In addition, patients with *TRANK1* rs71947865 (G) have more severe cognitive symptoms, a longer duration of 1^st^ episode, a more frequent family history of KLS, an older age of onset, and more behavioral disinhibition and altered speech during episodes, generally suggesting more severe disease (**Table 2**, full list of variables tested in **Dataset S4**). Types of birth difficulties are summarized in **Dataset S5**. In a further analysis of the association of clinical variables with the genotype dose of the *TRANK1* deletion, we adjusted for reports of birth difficulties. We noted that cognitive signs, age of onset and longer 1^st^ episode duration were significantly (p<0.05) increased in KLS cases with TRANK1 deletion allele, while there was a trend to association of TRANK1 deletion allele with current depression in KLS cases (**Table 3**).

**Table 2:**
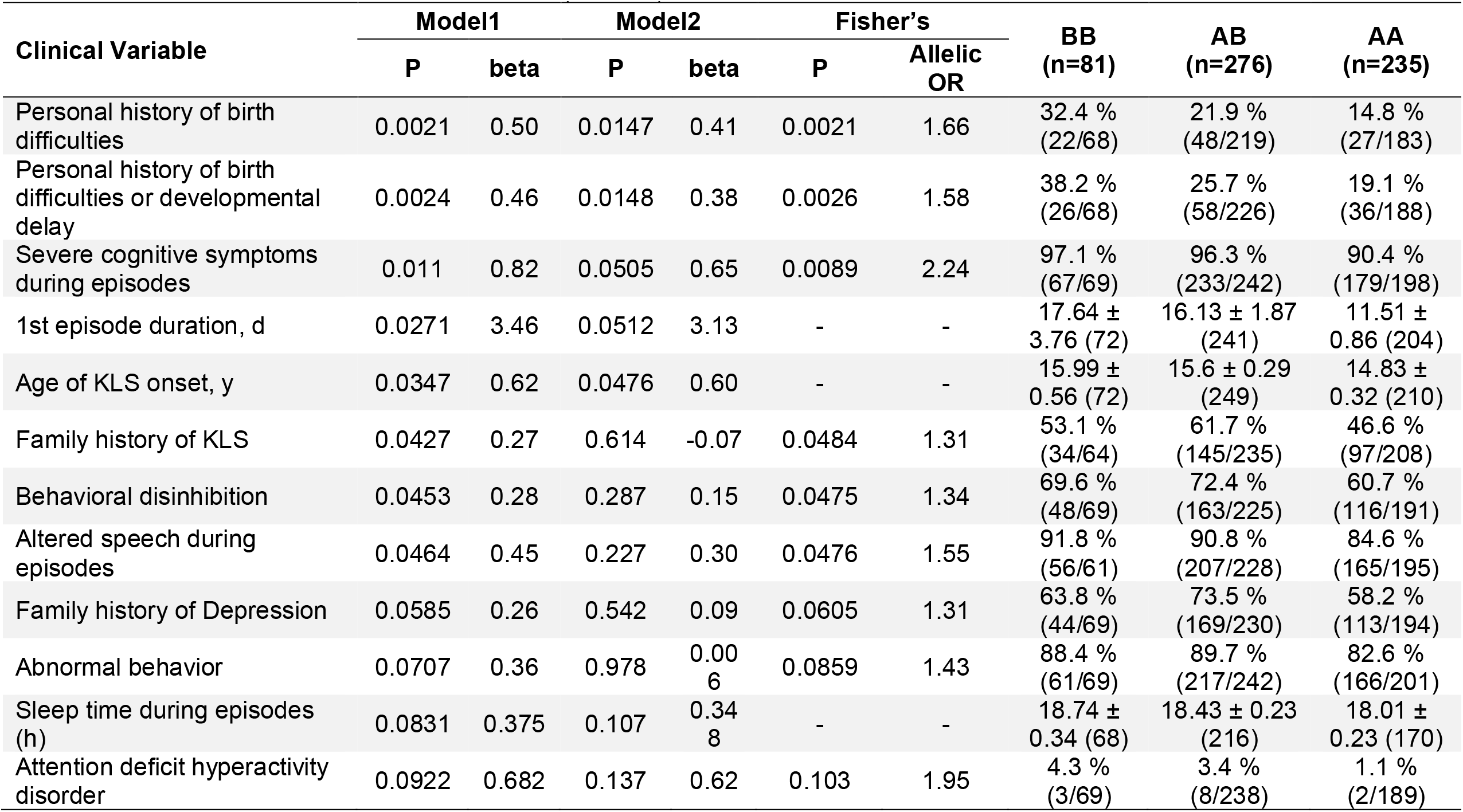
TRANK1 deletion (rs71947865) and its univariate association with KLS clinical variables. We coded the minor allele additively and used linear regression for continuous variables and logistic regression for categorical variables : Model1 - the minor allele genotype was fit as a function of the clinical variable, Model2 – was similar to Model1 except for including Ethnicity, Gender and Missing data proportion as three covariates. A Fisher’s exact test of allelic counts was further performed for categorical variables. In these analyses 592 KLS cases were included (Frequency % | mean ± sem was computed). BB-allele G/G, AB- GGGAGCCA/G, AA - GGGAGCCA/ GGGAGCCA, *Minor Allele G (deletion) was the effect allele.

| Clinical Variable | Model1 |  | Model2 |  | Fisher's |  | BB<br>(n=81) | AB<br>(n=276) | AA<br>(n=235) |
| --- | --- | --- | --- | --- | --- | --- | --- | --- | --- |
|  | P | beta | P | beta | P | Allelic<br>OR |  |  |  |
| Personal history of birth difficulties | 0.0021 | 0.50 | 0.0147 | 0.41 | 0.0021 | 1.66 | 32.4 %<br>(22/68) | 21.9 %<br>(48/219) | 14.8 %<br>(27/183) |
| Personal history of birth difficulties or developmental delay | 0.0024 | 0.46 | 0.0148 | 0.38 | 0.0026 | 1.58 | 38.2 %<br>(26/68) | 25.7 %<br>(58/226) | 19.1 %<br>(36/188) |
| Severe cognitive symptoms during episodes | 0.011 | 0.82 | 0.0505 | 0.65 | 0.0089 | 2.24 | 97.1 %<br>(67/69) | 96.3 %<br>(233/242) | 90.4 %<br>(179/198) |
| 1st episode duration, d | 0.0271 | 3.46 | 0.0512 | 3.13 | - | - | 17.64 ±<br>3.76 (72) | 16.13 ± 1.87<br>(241) | 11.51 ±<br>0.86 (204) |
| Age of KLS onset, y | 0.0347 | 0.62 | 0.0476 | 0.60 | - | - | 15.99 ±<br>0.56 (72) | 15.6 ± 0.29<br>(249) | 14.83 ±<br>0.32 (210) |
| Family history of KLS | 0.0427 | 0.27 | 0.614 | -0.07 | 0.0484 | 1.31 | 53.1 %<br>(34/64) | 61.7 %<br>(145/235) | 46.6 %<br>(97/208) |
| Behavioral disinhibition | 0.0453 | 0.28 | 0.287 | 0.15 | 0.0475 | 1.34 | 69.6 %<br>(48/69) | 72.4 %<br>(163/225) | 60.7 %<br>(116/191) |
| Altered speech during episodes | 0.0464 | 0.45 | 0.227 | 0.30 | 0.0476 | 1.55 | 91.8 %<br>(56/61) | 90.8 %<br>(207/228) | 84.6 %<br>(165/195) |
| Family history of Depression | 0.0585 | 0.26 | 0.542 | 0.09 | 0.0605 | 1.31 | 63.8 %<br>(44/69) | 73.5 %<br>(169/230) | 58.2 %<br>(113/194) |
| Abnormal behavior | 0.0707 | 0.36 | 0.978 | 0.00<br>6 | 0.0859 | 1.43 | 88.4 %<br>(61/69) | 89.7 %<br>(217/242) | 82.6 %<br>(166/201) |
| Sleep time during episodes (h) | 0.0831 | 0.375 | 0.107 | 0.34<br>8 | - | - | 18.74 ±<br>0.34 (68) | 18.43 ± 0.23<br>(216) | 18.01 ±<br>0.23 (170) |
| Attention deficit hyperactivity disorder | 0.0922 | 0.682 | 0.137 | 0.62 | 0.103 | 1.95 | 4.3 %<br>(3/69) | 3.4 %<br>(8/238) | 1.1 %<br>(2/189) |

**Table 3:** Association of clinical variables with genotype dose of TRANK1 deletion (rs71947865) after adjusting for medical history of birth difficulties, gender, referrer/race & missing data proportion in 592 KLS cases (linear regression for continuous variables and logistic regression for categorical variables). (Frequency % | mean ± sem was computed). BB- allele G, AB- GGGAGCCA/G, AA - GGGAGCCA/ GGGAGCCA, *Minor Allele G (deletion) was the effect allele.

| Clinical Variable | P-value | beta | BB<br>(n=81) | AB<br>(n=276) | AA<br>(n=235) |
| --- | --- | --- | --- | --- | --- |
| Severe cognitive symptoms during episodes | 0.011 | 0.99 | 97.1%(67/69) | 96.3%(233/242) | 90.4%(179/198) |
| Age at KLS onset, y | 0.026 | 0.76 | 15.99 ± 0.56 (72) | 15.6 ± 0.29 (249) | 14.83 ± 0.32 (210) |
| 1st episode duration, d | 0.046 | 3.63 | 17.64 ± 3.76 (72) | 16.13 ± 1.87 (241) | 11.51 ± 0.86 (204) |
| Current depression | 0.097 | 0.50 | 7.2%(5/69) | 7.1%(17/238) | 2.7%(5/188) |

**Table 4:** A gene set enrichment analysis was carried out on the combined discovery and follow-up cohort of KLS cases using the MAGMA module. A total of 5346 sets including curated gene sets, KEGG pathways, Reactome pathways and GO biological processes derived from msigdb6.1 were used. (*5% FDR P value was computed*). Beta is the direction of association of the pathway/geneset with the KLS case control status.

| Gene Set Name | Ngenes | Beta | P value | FDR P value |
| --- | --- | --- | --- | --- |
| Reactome_Rora_Activates_Circadian_Expression | 22 | 1.03 | 1.29x10<sup>-7</sup> | 0.00069 |
| Reactome_Circadian_Repression_Of_Expression_By_Rev_Erba | 21 | 0.97 | 2.17x10<sup>-6</sup> | 0.00579 |
| Go_Regulation_Of_Tumor_Necrosis_Factor_Superfamily_Cytokine_Production | 88 | 0.29 | 0.00050 | 0.61043 |
| Go_Positive_Regulation_Of_Multicellular_Organismal_Metabolic_Process | 17 | 0.66 | 0.00051 | 0.61043 |
| Reactome_Bmal1_Clock_Npas2_Activates_Circadian_Expression | 33 | 0.51 | 0.00057 | 0.61043 |

*TRANK1* genotyping was conducted in 195 controls with birth difficulty data available. These controls were from the KLS case control study reported by Arnulf et al. (3), with further addition of controls (friend or spouse of KLS patient) over more recent years. In the control sample, 22 subjects reported birth difficulties and, unlike in the KLS cases we found no significant association of birth difficulties with *TRANK1* rs71947865 genotype (p=0.5, OR=1.05). To formally test for an interaction effect of *TRANK1* genotype on birth difficulties in KLS versus controls, we used the combined KLS cases-control samples matched by PCA. Whereas both birth difficulties and *TRANK1* had a significant effect on KLS risk (p<0.05), the interaction term of TRANK1 genotype and presence of birth difficulties did not reach statistical significance. Note that the sample of controls with clinical (birth difficulty) data available is small and, although it was also used in the GWAS as a control, most of the controls used for GWAS were drawn from the Genetic Epidemiology Research on Aging (GERA)(33) cohort and/or other controls obtained through other studies from the same countries, which come from similar countries and collaborators.

### The TRANK1 association is birth year dependent

As indicated above, our analysis revealed a significant association of the *TRANK1* variant with birth difficulties in KLS patients but not controls. As perinatal care has improved over the last decades (34-39); https://www.cdc.gov/nchs/data/nvsr/nvsr56/nvsr56_10.pdf; https://mchb.hrsa.gov/chusa14/health-status-behaviors/infants/infant-mortality.html), most notably in the 1990-2000s, we asked if the year of birth was an essential factor correlated to KLS in conjunction with *TRANK1* rs71947865 variant. A possible birth year dependent *TRANK1* interaction is also supported by the recent largest bipolar disorder GWAS ever published, where it was noteworthy that the strong *TRANK1* SNP association found in the discovery sample in bipolar disorder cases did not replicate in a new, presumably more recent, bipolar GWAS replication set (15), a caveat being that birth year data was not made available in this study (15) for all but one replication cohort where the birth year range was 1981-2005. Further, later reported *TRANK1* ORs for bipolar disorder decreased in size from earlier studies in Brazil (40) and other countries (41-43) to the most recent metanalysis (15), even independently of the non-significant replication sample of this most recent metanalysis (15). This is illustrated in **Dataset S3**, where published TRANK1 association results are reported, with the caveat that the same samples are typically used cumulatively across studies.

In a subset of the full cohort (discovery and replication sets) when a valid birth year was available (n=650), we performed *TRANK1* SNP association by birth year stratified into three approximately equal-sized groups; 1987 or earlier (n=220, median year of birth: 1978), 1988-1994 (n=202, median year of birth: 1992) and 1995-2000’s (n=228, median year of birth: 1999). Using genetic principle component matched controls for each stratum mainly derived from the GERA cohort (birth year range ∼1928 to 1989), we observed a decrease in *TRANK1* SNP rs71947865 association with time (**Figure 3A** and **Dataset S6A**). Notably, while individuals born in 1987 or earlier had the strongest association *TRANK1* association (p=8.3×10^−4^; OR=1.41), this effect decreased in KLS cases born 1988-1994 (p=7.48.3×10^−3^; OR=1.34) to finally disappear in the 1995-2000’s group (p=0.31; OR=1.11). In addition, within KLS cases, a Fisher’s test of allelic counts was performed, birth year bin 1987 or earlier was compared to 1988-1994 and 1995-2000’s in 2×2 contingency table. We observed highest association of *TRANK1* SNP rs71947865 (p=0.002) in year bin 1987 or earlier when compared to the minor allele count of *TRANK1* SNP rs71947865 in the birth year bin 1995-2000’s (see **Dataset S6B**).

**Figure 3:**
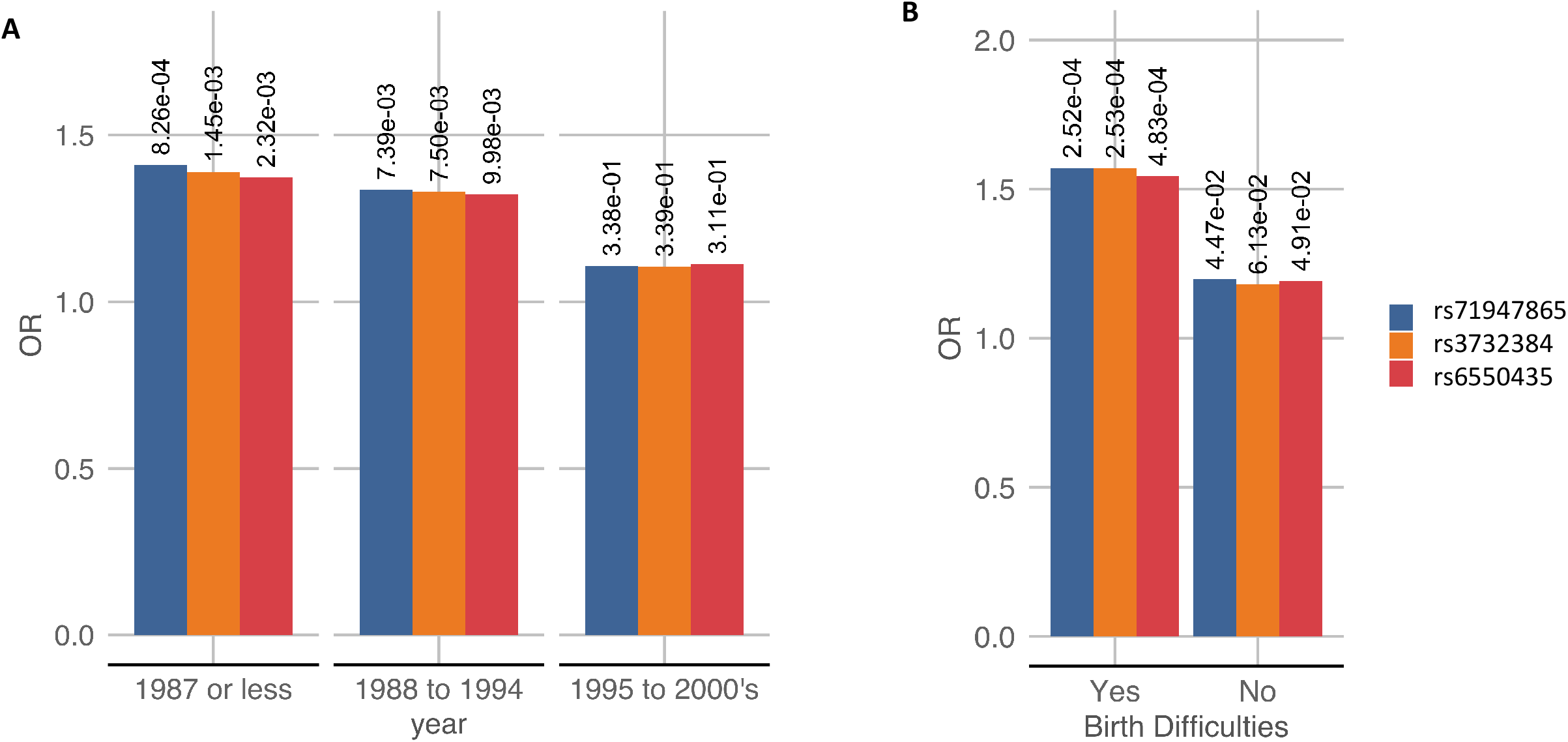
**(A)**The *TRANK1* region SNP associations in KLS cases stratified by birth years, the y-axis represents the odds ratio of the effect allele while the x-axis is the birth year. Inset P value of the association is labelled. **(B)** The *TRANK1* region SNP associations in KLS cases stratified by birth difficulties, the y-axis represents the odds ratio of the effect allele while the x-axis is the presence of birth difficulties.

### GWAS Analysis of follow up KLS cases with and without reports of birth difficulties

To replicate these findings, we collected and genotyped 171 new KLS cases collected between 2017-2019, collecting information on birth difficulties whenever possible. In the resulting sample set, birth difficulties data were available for 151 of the 171 cases. In the overall replication sample (n=171), generally born more recently (median year of birth: 1998, Range: 1960 to 2015), TRANK1 (rs71947865) was not significantly associated with KLS (p=0.1, OR=1.1) (**Table 1** and **Dataset S2**). Strikingly, however, when we stratified this sample into cases with versus without birth difficulties, then a significant *TRANK1* association was seen only in the first group (p=0.01; OR=1.54 in n=59 cases reporting birth difficulties, **Dataset S7**). In KLS cases with no birth difficulties, little effect was observed (p=0.4; OR=1.12 in n=88 cases). No birth information was available for n=24 patients. A combined *TRANK1* SNP meta-analysis (OR=1.4;p=3.5×10^−8^) of both discovery and replication cohorts is presented in **Table 1**

Our prior analyses of the discovery cohort had revealed significant association of *TRANK1* variant rs71947865 with birth difficulties, we therefore determined the association of *TRANK1* rs71947865 variants in KLS cases with birth difficulties when compared to controls individuals in both discovery and replication cohorts. We observed that *TRANK1* rs71947865 variants were significantly associated birth difficulties in KLS cases from the both discovery cohort (p=0.002; OR=1.61, see **Dataset S8A**) and the follow-up sample of KLS cases (p=0.03; OR=1.53). A combined analysis of KLS cases with and without birth difficulties from combining both discovery and replication cohort compared to genetic PCA matched controls also revealed significantly increased TRANK1 variant rs71947865 frequencies in KLS cases with birth difficulties (Fisher’s test, OR=1.54; p=4.83×10^−4^, **Figure 3B & Dataset S8B**) while in KLS cases without birth difficulties, the effect was weaker (Fisher’s test, OR=1.20;P=0.04).

### Fine mapping of the KLS signal across ethnicity

GWAS studies in schizophrenia and bipolar disorders have mapped the TRANK1 signal at SNP rs9834970 (r^2^=0.74 with KLS SNP rs71947865) in Caucasians (15, 42-44), while when a mixed ethnic sample that included Asians and Caucasian was used, the top SNP is rs3732386 (16, 45-47), a signal entirely linked (r^2^=1) with the KLS associated SNP rs71947865 (see **Dataset S3**; **SI Appendix, fig. S2A-sC**).

Since our KLS sample is derived from multiple ethnicities, as an additional validation, we sought to determine *TRANK1* loci association KLS cases of European/Caucasian origin only. In the overall European KLS meta-analysis, we observed that the *TRANK1* region SNP rs71947865 (OR=1.28; p=1.9×10^−4^) was retained as the top association (see **Dataset S9** and **SI Appendix, fig. S2A-C**) with no evidence of heterogeneity. Our KLS European/Caucasians only analysis (discovery plus replication cohort) revealed that rs9834970 was among the top associations in the loci (**SI Appendix, fig. S2D** and **Dataset S9**). The rs3732386 and rs71947865 were the top associations in the *TRANK1* region and were dependent on the existence of associated birth difficulties (**SI Appendix, fig. S2**). Further, on comparing the associations in the TRANK1 loci in discovery KLS versus combined schizophrenia and bipolar disorder GWAS, there is remarkable overlap of signals with only the magnitude of ORs being 1.3x times increased in KLS (see **SI Appendix, fig. S2D**).

### Replication of Polygenic risk score of KLS

We found significant evidence that the KLS discovery cohort polygenic risk scores (PRS) were indicative of increased genetic liability to KLS phenotype in a follow-up replication cohort (n=171) under various p-value thresholds using PRSice2 (48). The most predictive p-value threshold was 0.5 (*pseudo R2* p=2×10^−22^; **Figure 4**) using 81,570 variants. P-value thresholds ranging from 0.05 to 0.4 were also indicative of increased genetic liability to KLS (p<1×10^−20^, mean *pseudo R2 =0*.*14, see* **Dataset S10**). Further, as an additional control, we also used only Caucasian KLS sample discovery meta-analysis (n=299) to build PRS with validation only in Caucasian KLS replication sample (n=160), the PRSs were indicative again of increased genetic liability to KLS (*pseudo R2 =0*.*094) with* the best score at p=0.4 threshold (see **SI Appendix, fig. S3**).

**Figure 4:**
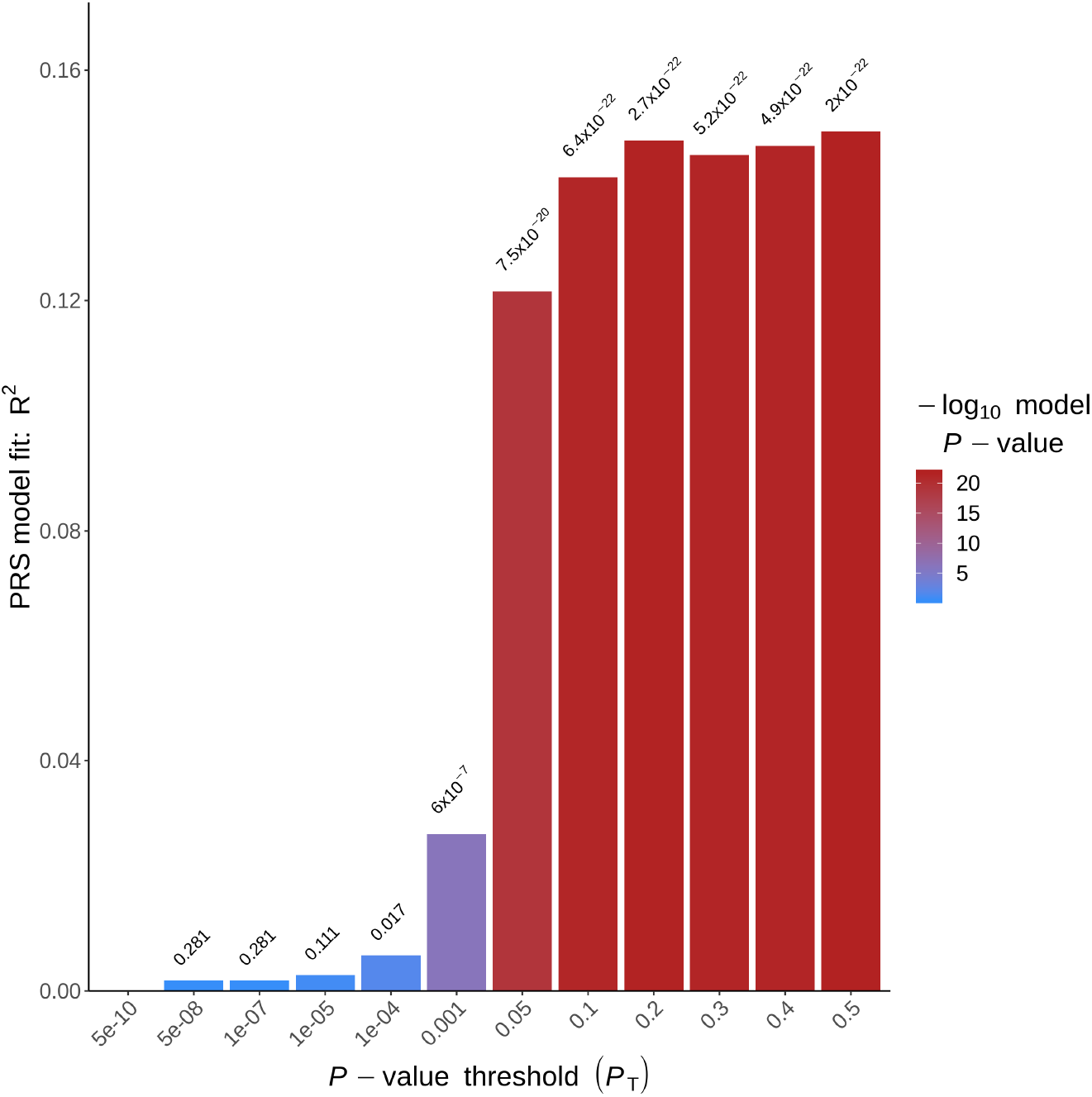
Variance explained by PRS (polygenic risk scores) built on the KLS discovery cohort when used to predict on the replication cohort, the y-axis is the % variance explained by the PRS while the x-axis represents the p-value thresholds.

Considering the results above, we further built PRSs for Bipolar disorder (15), Schizophrenia (49), circadian chronotype (50), sleep duration (51), and excessive daytime sleepiness (52) using previously published GWAS studies, and evaluated their predictive performance on our KLS follow up cohort. Shared KLS genetic liability (*pseudo R2 =0*.*06)* was found using bipolar disorder and schizophrenia GWAS summary statistics as the training set (see **SI Appendix, fig. S3** and **Dataset S10**). LD score regression analysis estimated the total observed scale heritability h2=0.09 (intercept=1.044) and also revealed significant genetic overlap of KLS with bipolar disorder (rg=0.41;p<0.05) and depressive symptoms (rg=0.32;p<0.05, **Dataset S11**).

### Gene Pathway analysis

Biological processes pathway enrichment analysis was next conducted using the MAGMA (53). The MAGMA module gene-based association analysis was first performed in the combined discovery-replication sample summary statistics mapping SNPs to genes with 1kb window upstream and downstream. This first step entails collapsing mean individual SNP statistics into a per -gene level statistic. Then a gene p-value is obtained by using a known approximation of the sampling distribution as detailed in MAGMA (53). The gene-based p values converted to z-scores are then used to conduct gene-set enrichment analyses on gene sets derived from MsigDBv6.1 (5346 curated gene sets). At 5% FDR (false discovery rate), pathways “*Reactome-RORA activates circadian expression*”(54) and “*Reactome-Circadian Repression of Expression By Rev ERBA*”(55) were observed as significantly enriched in KLS genetic association analyses (see **Table 4**). Notably these 2 pathways shared identical gene members except for single gene. Gene based association tests within these enriched pathways are presented in **Dataset S12**. Both ROR Alpha (retinoic acid receptor-related alpha) and Rev ERBA are critical transcriptional regulators of periodic expression of BMAl1 gene, directly regulating circadian pathways (56-59). Other circadian pathways of interest included “*Reactome-Bmal1 Clock NPAS2 Activates Circadian Expression*” but did not reach 5% FDR threshold.

### KLS Exome sequencing analysis

Targeted exome sequencing was performed in 32 individuals with KLS derived from 16 multiplex families (see **Dataset S13A**). The variant frequencies and variant by variant analyses compared to 286 European controls from 1000 genomes phase 3 were largely unremarkable. Damaging or probably damaging variants in each gene were collapsed and mutational burden in KLS probands and/or shared by at least 3 families were identified and presented below.

### LMOD3 gene variants are not associated with KLS

Following up on a recent study suggesting increased mutation load in the LMOD3 gene in familial KLS cases (60), we analyzed the LMOD3 burden in our cohort of KLS probands and sporadic cases (n=17) compared to controls (n=286). We observed no significant differences in mutational burden in LMOD3 gene between cases and controls (p=0.13; see **Dataset S13B**). Variant level analyses in KLS probands and sporadic cases (n=17) revealed a previously reported single site rs35740823 (chr3:69171290; p.R83H) to be enriched in KLS cases (allelic OR=3.8; p=0.06) as compared to controls (60) (see **Dataset S13B**). We therefore imputed this LMOD3 variant rs35740823 and analyzed for associations in 151 KLS cases and 478 controls derived from replication cohort and predominantly of European origin but did not find any remarkable differences (OR=1.1; p=0.77; **Dataset S13B**).

### Gene mutational burden test in KLS

We also compared the overall frequency of modifier variants tagged using SnpEff (61) as moderate or high effects in all genes in all familial KLS cases versus 1000 genome controls (n=286) running gene based mutational burden analyses (**Dataset S14A**), matched for ethnicity and SNP principal components. Although significant effects were found, none were found in genes within the *TRANK1* loci. We however observed increased mutational burden in the rare alleles in genes NRXN2, *GABRG2, CHORDC1*, AATK and *CASP7* genes (FDR p<1×10^−4^) among probands (n=17) relative to 1000 genomes reference samples (**Dataset S14A**). Genes with excessive rare alleles across each KLS family were next analyzed. Notably, GABRG2, CASP7 and ANKRD31 among other genes were shared across at least 3 KLS families (see **Dataset S14B**).

Further, pathway burden test from the list of 21 genes belonging to the circadian regulation pathway identified through the MAGMA analysis revealed significantly increased minor allele burden (see **Dataset S15A**) in the whole KLS cohort (n=32) compared to European origin controls. We also used pathway sets from msigDB6.1 to profile mutational burden in each pathway in KLS probands (n=14). Notably, we found increased mutational burden in GO GAMMA AMINOBUTYRIC ACID SIGNALING PATHWAY (FDR p=0.007) in KLS probands among other pathways that included many neuronal pathways (see **Dataset S15B**). This agrees with results of the gene burden test where we observed significantly increased burden in GABRG2 in KLS probands.

## Discussion

Our study is the first to explore the interaction of genetic and environmental contributions to the pathophysiology of KLS. Results of GWAS revealed that TRANK1 gene variants were predisposing and associated with birth difficulties in KLS. PRS constructed on discovery GWAS analysis found strongly replicable PRS that strikingly overlapped with bipolar disorder and schizophrenia. Pathway analysis also revealed the involvement of behavioral and circadian rhythmicity genes.

This large study did not confirm findings made in other smaller genetic studies of KLS. For example, a 20-year-old study had suggested an HLA-DQ2 association (12) in KLS, but this was not confirmed either in other studies (3, 8, 13) nor in our current analyses. A recent study (60) focused on a large Saudi pedigree KLS affected family where a rs111848977 p.E142D mutation in *LMOD3* was observed to cosegregate. This study also reported an increased burden of damaging mutations, including p.R23H in LMOD3 in KLS, a gene involved in nemaline myopathy 10 (62). In our sample of 32 familial cases, we could not find any overall increase in the frequency of *LMOD3* mutations and, although p.R83H was nominally increased in familial cases, we did not find this association in a larger cohort of 151 KLS cases (see **Dataset S13B**).

Although the phenomenology of KLS is quite different from that of bipolar disorder or schizophrenia, these syndromes have some clinical similarities. Most notably, in both KLS and bipolar disorder, patients are asymptomatic between episodes. Similarly, exacerbations are usually episodic in some schizophrenia. In KLS, however, onset/offset of episodes is rapid (typically within one day), while it is generally slower in most bipolar patients, except for “rapid cyclers” (63). Further, although not a dominant feature, depression, anxiety, or hallucinations/delusions can occur during KLS episodes (3, 4, 6, 64) and at the end of episodes, some patients may have brief (1-2 days) period of elation, insomnia, and logorrhea which can resemble a maniac mini-episode.

Most interestingly, hypersomnia is a common feature of depressive episodes in bipolar type 1 disorder (65-67), unlike in other depressions where patients most often exhibit insomnia. Like KLS, bipolar disorder is also associated with cognitive deficiencies, although these are milder (68, 69) as compared to KLS patients. This, together with the fact that lithium is the only treatment reported to have an effect in preventing KLS episodes (7), suggests possible shared pathophysiology with bipolar disorder. Pathophysiological overlap among KLS, bipolar and schizophrenia is also in line with genetic studies in the United Kingdom (UK) Biobank cohort that have shown that the genetic architecture of long self-reported sleep duration overlaps with that of bipolar disorder, in contrast with genetic architecture of insomnia that shares similarity with anxiety and Restless Leg Syndrome (70). Most strikingly, strong correlations between PRS scores of long sleep duration and bipolar disorder type 1 in the UK Biobank sample was found (71). This suggests that hypersomnia, periodic or not, could share some commonality with bipolar disorder and schizophrenia.

One possible hypothesis that KLS, like bipolar disorder and other psychosis, is characterized by periodic instability of specific brain networks involving, in the case of KLS, reduced oxygen flow in cortical and subcortical areas, as reported by imaging studies(72). In this syndrome, cognitive and derealization symptoms may correlate with decreased activity in cortical areas, whereas decreased activity in thalamus and hypothalamus may be involved in the functional hypersomnia observed in disorder (72-74). Involvement of circadian or behavioral rhythmicity genes may be essential for maintaining hypersomnia episodes once an episode is initiated, for example, if subjects are suddenly spending much time in bed, unexposed to light, a behavior which could result in difficulties entraining in genetically susceptible individuals (75). This hypothesis could also explain why patients with KLS can exhibit Sleep Onset REM Periods during MSLTs (the circadian clock strongly drives occurrence of REM sleep) (76, 77), a feature also associated with shift work (78). Other investigators have found abnormal circadian entrainment during KLS episodes using actigraphy (79). A role for circadian abnormalities has also been suggested in bipolar disorder type 1 (67, 80, 81). Subjects with bipolar type 1 also reported increased eveningness tendencies (71), although this was not found in patients with KLS (3).

The most striking finding of this study is the observation of a strong genome-wide significant association of KLS with *TRANK1*-region variants [OR∼1.50] that are strongly associated with bipolar disorder [OR∼1.12] (43, 82, 83) and schizophrenia [OR∼1.08] (46, 84-86), but not mood instability (87). Although it remains possible this effect could be due to a rarer variant linked with this haplotype in KLS, this is more likely mediated by a similar effect of these common variants across these diseases. Of notable importance is the fact the *TRANK1* effect size in this study is 4-5 orders of magnitude higher (OR∼1.5) than in studies in these other pathologies (OR∼1.1), explaining detection of association in a small sample of less than 1,000 KLS patients. This association with KLS is thus unlikely to be explained by contamination of our current KLS cohort with bipolar cases, although the opposite, contamination of bipolar or schizophrenia samples with KLS patients could explain weaker associations in these other pathologies.

The reason for the weaker than expected *TRANK1* replication in the KLS replication cohort became evident after stratifying our discovery sample by birth decade, finding that the *TRANK1* association was highest in subjects born before the 1980s and decreased thereafter. Further, KLS subjects with *TRANK1* polymorphism have reported increased birth difficulties (summarized in **Dataset S5**). We hypothesize that improved perinatal outcomes that have occurred in the past 50 years may explain this tendency (34-39); https://www.cdc.gov/nchs/data/nvsr/nvsr56/nvsr56_10.pdf; https://mchb.hrsa.gov/chusa14/health-status-behaviors/infants/infant-mortality.html). The *TRANK1* birth difficulties relationship was also confirmed in the independent replication sample, where we found that only the subset of KLS patient reporting birth difficulties had a significant *TRANK1* association. Overall, patients who reported birth difficulties had the highest association (Fisher’s test, OR=1.57; p=2.5×10^−4^) both in the discovery and replication cohorts (**Dataset S9A-B**). *TRANK1* association also increases disease severity during episodes independently of birth difficulty effects (**Table 3**).

Interestingly, although the association with *TRANK1* was the highest genome significant finding in bipolar disorder in a recent metanalysis of 32 cohorts from 14 countries in Europe, North America and Australia, totaling 20,352 cases and 31,358 controls of European descent (rs9834970; OR=1.10, p=5.5 × 10^−14^) (15), but, as in our current KLS sample, replication in follow-up bipolar disorder samples totaling 9,412 cases and 137,760 controls of European ancestry did not replicate (OR=1.02, p=0.15). This finding deviated more than expected by chance (p=0.01, although not significant if Bonferroni corrected for all loci detected in the study). We hypothesize a similar time dependent *TRANK1* birth difficulty interaction may play a role in bipolar disorder and schizophrenia. Indeed, these disorders are known to be strongly associated with early life complications (17-28, 88, 89), and data in the UK Biobank cohort indicates a dose-dependent increased prevalence of major depressive disorders and bipolar disorders in association with reported low birth weight (90), even when controlled for a history of the physical disorder known to be associated with low birth weight.

Two possibilities, which are not necessarily mutually exclusive, could explain this data. First, *TRANK1* region genes could affect placental physiology, modulating the risk of birth difficulties and long-term fetal damage that could manifest as KLS or other disorders. Ursini et al. (31) found strong significant genetic interaction between the PRS of schizophrenia and the occurrence of early life complications in multiple case-control samples. Further, *MLH1, LRRFIP2*, and *GOLGA4* associated with these overall genetic components were highly expressed in the placenta and modulated by pathological placental processes such as preeclampsia and intrauterine growth restriction but not in other disease processes (31). Problematically, however, recent papers did not observe that the KLS associated SNPs identified in this study had substantial effects on placental expression of these genes (31, 32, 91, 92). Further, *TRANK1-*region polymorphisms are not known maternal or fetal genetic factors associated with low birth weight (93-95), prematurity (96), or birth complications (91, 97) in large samples, and were not associated with birth difficulties in a small sample of 198 controls similarly ascertained to our KLS subjects for birth difficulties. Finally, this hypothesis would not explain the time dependency/cohort effect of the *TRANK1* association we observed with KLS, which is likely due to improved or changes in perinatal care with time.

Another possible explanation, which we favor as it better fits our results and the lack of effects of TRANK1 on birth difficulties in previously published studies, is that newborns carrying *TRANK1*-region polymorphisms are more susceptible to injury in the presence of birth difficulties (notably hypoxia) in a way that produces behavioral instability such as KLS or other psychiatric conditions such as bipolar disorder in adulthood. This would be a true gene x environment interaction, although our study lacked sufficient control to formally test this hypothesis statistically. The mechanism underlying this phenomenon, which could have evolutionary advantages in times of stress (98), remains to be investigated. Interestingly, rs9834970, an SNP linked with the *TRANK1* region susceptibility haplotype is associated with decreased expression of *TRANK1* in iPSCs and neural progenitor cells (99); such effect has not been reported in the adult brain, but, as discussed, *TRANK1* associated polymorphism reduces expression in various dividing tissues such as blood, esophageal mucosa, and B cells. The iPSC/neuronal precursor cell effect was rescued by valproic acid but not lithium, and the TRANK1 gene knockdown was found to modulate the expression of many genes, suggesting developmental effects (99).

Because the susceptibility haplotype modulates expression of many other genes in a tissue-specific manner, however, we believe the study of other genes in this region across tissues and development with the susceptibility haplotype will be necessary before concluding that *TRANK1* is the gene behind this genetic association which is a limitation of our analyses. Additional studies aiming at studying more precisely which types of birth difficulties are associated with *TRANK1* in these disorders is also needed. Our results offer new clues on the developmental window in which the *TRANK1*-susceptibility haplotype could affect risk for KLS and mental disorders in adulthood and mandates a more careful analysis of what and how the reported birth difficulties interact with specific *TRANK1* genetic variants.

Our data support a genetic predisposition in the presence of *TRANK1* polymorphisms and links KLS, behavioral rhythmicity, and bipolar disorder. Polygenic risk scores derived from discovery KLS samples are strongly predictive of the KLS phenotype in the replication sample. Further, PRSs support a genetic overlap with bipolar disorder and schizophrenia with the caveat being that these associations in various p thresholds were unadjusted for multiple comparisons and likely are underpowered since only replication follow up KLS sample was used. Strikingly, the temporal effect of *TRANK1* polymorphism was observed in KLS cases in relation to the year of birth. In the presence of birth difficulties, the *TRANK1* polymorphism may predispose to pathophysiological consequences such as KLS, bipolar disorder, and schizophrenia.

## Materials and Methods

### Inclusion criteria for KLS and origin of cases

Patients with KLS were included if they met International Classification of Sleep Disorders (ICSD3) criteria for the disorder including the presence of recurrent episodes (≥2) of severe hypersomnia (>18/24h) lasting days to weeks, associated with cognitive disturbances (confusion, amnesia, derealization), behavioral abnormalities (major apathy, megaphagia, and hypersexuality). These episodes had to be interspersed by long periods of normal sleep, cognition, behavior, and mood. A sleep-trained physician-diagnosed all patients. They were also asked to answer a detailed questionnaire, or a systematic interview as described (3, 6). Informed consent, in accordance with governing institutions, was obtained from all subjects. The research protocol was approved by Institutional Review Board (IRB) panels on Medical Human Subjects at Stanford University, and by respective IRB panels in each country providing samples for the study.

### KLS cases

The discovery sample of 673 KLS cases includes 261 cases recruited in the United States (US), with the help of the Kleine Levin Syndrome Foundation (KLSF). US patients were diagnosed by an outside physician or referred by the KLSF (Steve Maier, Neal Farber), and the diagnosis confirmed at Stanford, where questionnaire and blood sample was collected. Other major recruitment sites include Pitié-Salpêtrière, Paris, France (Dr. Isabelle Arnulf, n=140), University of Peking, Beijing, People’s Republic of China, (Dr. Fang Han, n=62), Taiwan University, Taipei, Republic of China, Taiwan (Dr. Yu-Shu Huang, n=58), Guy de Chauliac Hospital, Montpellier, France (Dr. Yves Dauvilliers, n=35), Safra Children’s Hospital, Sheba Medical Center, Tel Aviv University, Tel Aviv, Israel (Dr. Yakov Sivan, n=34), Hepata Clinic, Germany (Dr. Gert Mayer, n=24), Uppsala University, Sweden (Dr. Anne-Marie Landtblom, n= 15), Hospital Robert Debre, Paris, France(Dr. Michel Lecendreux, n=7), Tokyo Metropolitan Institute of Medical Science Tokyo, Japan (Dr. Makoto Honda, n=7),University of Bologna, Italy (Drs Fabio Pizza and Guiseppe Plazzi, n=6), Maynei Hayeshua Medical Center, Bnei Barak, and The Sackler Faculty of Medicine, Tel-Aviv University (Dr. Natan Gadoth, n=6), Akita University, Akita, Japan (Dr. Kanbayashi, n=2), St. Vincent’s Hospital,The Catholic University of Korea (Dr. Seung Chul Hong, n=2), 1st Faculty of Medicine and General Teaching Hospital, Prague, Czech Republic (Dr. Sona Nevsimalova, n=2), Bambino Gesù Children Hospital, Rome, Italy (Dr. Marina Macchialio, n=2) and other centers who contributed single sample (n=10).

The replication cohort included 171 additional KLS cases. These included cases from Pitié-Salpêtrière, Paris, France (Dr. Isabelle Arnulf, Dr. Smaranda Leu-Semenescu and Dr. Pauline Dodet, n=66), Uppsala University, Sweden (Dr. Anne-Marie Landtblom, n= 26), Kleine Levin Syndrome Foundation (n=23), University of Bologna, Italy (Drs Fabio Pizza and Guiseppe Plazzi, n=13), London Bridge Hospital, London, UK (Dr. Guy Leschziner n=11), Guy de Chauliac Hospital, Montpellier, France (Dr. Yves Dauvilliers, n=9), Emory University, Atlanta (Dr. David Rye n=6), Hepata Clinic, Germany (Dr. Gert Mayer, n=5), Stanford University, (Dr. Mignot, n=5), Tokyo Metropolitan Institute of Medical Science, Tokyo, Japan (Dr. Makoto Honda n =4) and other centers (n=3).

### GWAS patients and matched controls

Because the samples were collected over a long-time frame (2003-2019), genotyping platforms evolved, and hence each platform required its cohort of cases and controls **(Dataset S1)**. As a result, the discovery sample (n=673, 2003-2017) was subdivided into seven subcohorts of cases and controls based on genotyping platform/ethnicity differences. Cases and ethnically matched controls (n=15,339) were genotyped at UCSF genomics core with Affymetrix Axiom^®^ Genome-Wide LAT, EUR, CHB, Axiom^®^ 55066 arrays, Axiom^®^ world arrays, while multiethnic Axiom^®^ PMRA arrays were genotyped at Akesogen inc. These include three European ancestry cohorts, an Asian cohort, a Latino cohort, and Ashkenazi and Non-Ashkenazi Jewish cohorts. The replication sample (n=171, collected between the years 2017-2019), subcohort 8, was treated as a single sample and genotyped solely on Axiom^®^ PMRA arrays. DNA was extracted using Qiagen DNeasy kit, and samples were stored at -20c until genotyping, a minimum of ∼200ng was used for genotyping.

### Genotype analysis and imputation

Raw CEL files were imported into Affymetrix Genotyping console, and Affymetrix best practices workflow was followed. Genotype calls were exported as plink ped/map files (100). For CEL files generated from Axiom^®^ world arrays, primarily AffyPipe was used to call the genotypes and exported as plink ped/map files (101). Stringent QC was applied and variants and individuals with >0.05 missingness excluded, related individuals with identity by descent pihat>0.2 excluded and markers with Hardy-Weinberg equilibrium p<1e^-6^ in controls excluded. HapMap and 1000 genomes phase 3 reference population genotypes(102) were used to guide ethnicity matching in our sample sub cohorts. Population covariates were extracted using plink and visually inspected for unexpected deviations and outliers, after which Euclidean distance was used to accurately and automatically match cases and controls. Post QC, genotypes were phased one chromosome at a time with shapeit (103, 104) using the build37 coordinate system of the human genome. Phased haplotypes were then subject to imputation to 1000 Genomes phase 1 in 1mb chunks for each chromosome using impute2 (105).

### Association testing and variant prioritization

Frequentist association tests were then performed on the imputed genotypes by including genetic principal components (PCA) and missing data proportion using snptest (106). Meta-analysis was subsequently performed using cohort specific β estimates using a fixed effects model(106). The genome-wide significance threshold was set at p<5e-8 based on Bonferroni adjustment. Gene-based association tests, including pathway analysis were performed with MAGMA modules (107). Manhattan plots were constructed with R package qqman (108). Additional eQTL annotation was carried out using SNIPA, webserver (109), and GTEx v8 (30). eCAVIAR was used to perform eQTL colocalization (110). LD hub was used to calculate LD score regression and find shared SNP heritability across phenotypes (111). Polygenic risk scores were computed using PRSice2 (48), briefly imputed genotypes in BGEN format from the replication data set were quality controlled (maf >0.05 and impute info score >=0.9) and clumped for linkage disequilibrium (250 kb window; r2 = 0.1) and used as target dataset along with first five genetic PCAs. The common variants remaining after clumping from the base and target dataset were 104626. Apart from using summary statistics from the KLS discovery, we also used summary statistics from various other phenotypes. PRSs at various p-value thresholds (5×10^−10^,5×10^−8^,1×10^−7^,1×10^−5^, 0.0001, 0.001, 0.05, 0.1, 0.2, 0.3, 0.4, 0.5 and 1) were derived and validated in KLS replication cohort (n=171).

### Exome sequencing sample and matched controls

Agilent Sure Select V4 system workflow was used to capture regions of interest; the resulting fragments were subject to Illumina paired-end sequencing run with an average depth of 130x and average read length of 100 in the target regions for the KLS family DNA samples. The KLS family DNA consisted of 32 individuals derived from 16 multiplex families (see **Dataset S13A**). On the other hand, for controls, exome FASTQ files from 286 individuals part of 1000 genomes (predominantly CEU, TSI, GBR, IBS & FIN) were used. The FASTQ files were aligned to UCSC hg19 reference FASTA using BWA (112) and read groups inserted for downstream variant quality score recalibration. GATK best practices workflow (https://software.broadinstitute.org/gatk/best-practices/) was implemented, and joint genotyping performed on both cases and controls simultaneously. Raw variant call sets were subject to variant quality score recalibration, and variants passing the filters were then imported into plinkseq (https://atgu.mgh.harvard.edu/plinkseq/) and used in downstream association analysis. Annovar and SnpEff were used to annotate variants (61, 113). Following which moderate and high modifier variants were collapsed and gene-based burden tests in various sub-cohorts were carried out using plinkseq.

A gene burden test was carried out within each family, probands only and all the 32 individuals. Each analysis used the 1000 genomes European dataset matched by principle components as a control cohort. The burden analyses were focused only in rare variants (minor allele counts 2-20) and restricted to predicted to be moderate or high modifiers as tagged by Annovar and SnpEff. Further, genes part of the rhythmic behavior pathway were collapsed, and SKAT based gene burden tests were performed (114) with resulting p-values adjusted using a 5% false discovery rate (FDR).

### Taqman genotyping

To validate the genotype calls from SNP genotyping arrays, we conducted Taqman assays for *TRANK1* SNPs rs71947865 and rs6550435 on a random subset of KLS cases and controls typed across multiple platforms (n=122). Allele concordance for rs71947865 was 100%, while for rs6550435, it was 97% in 122 subjects.

### Clinical Associations

Demographic, and various clinical features within the KLS cohort (n=592) were analyzed for associations with *TRANK1* rs71947865 variant (deletion). Three kinds of analyses were performed, model1; A linear additive model was fit as a function of the clinical variable, Model2; same as model1 except for including covariates such as ethnicity/referrer code (geographical location) and missing clinical data proportion per individual, Fisher’s exact test of allelic counts was performed for all categorical clinical variables.

### Birth Year Associations

Two analysis strategies were pursued, first, when birth year was available for KLS cases (n=650), we stratified the KLS cases into three approximately equal-sized groups; 1987 or before (n=220, median year of birth: 1978), 1988-1994 (n=202, median year of birth: 1992) and 1995-2000’s (n=228, median year of birth: 1999). Within each group, we matched control samples based on genetic principle components. Fisher’s exact and chi-square tests on *TRANK1* rs71947865 allelic counts were performed to find significant difference in allele frequencies. A Fisher’s test of TRANK1 SNPs allelic counts was performed within KLS cases stratified by the 3 birth year bins, the minor allele counts in birth year bin 1987 or earlier was compared to minor allele counts 1988-1994 and 1995-2000’s in 2×2 contingency table.

## Data Availability

All genotyping calls can be requested by contacting the corresponding author.

## Data availability

All genotyping calls can be requested by contacting the corresponding author.

## Acknowledgments

We thank countless staff members, clinicians, patients, and patient advocates who are not listed here and have provided material and logistic support over decades for these 15 yearlong studies. The study was primarily funded by the Kleine-Levin Syndrome Foundation (KLSF, 2003-2020) and by NIH 1R01MH080957 (2007-2013) to EM. The French Kleine-Levin Syndrome research program is financed by a grant to IA (PHRC 070138) from the French Ministry of Health. HMO has been supported by the Academy of Finland (#309643), Yrjö Jahnsson Foundation and Oskar Öflund Foundation. Computing for this project was performed on the Stanford Genomics cluster supported by NIH 1S10OD023452-01. We would like to thank the Stanford Research Computing Center for providing computational resources and support that contributed to these research results.

## Supplementary Information

**Fig. S1.**
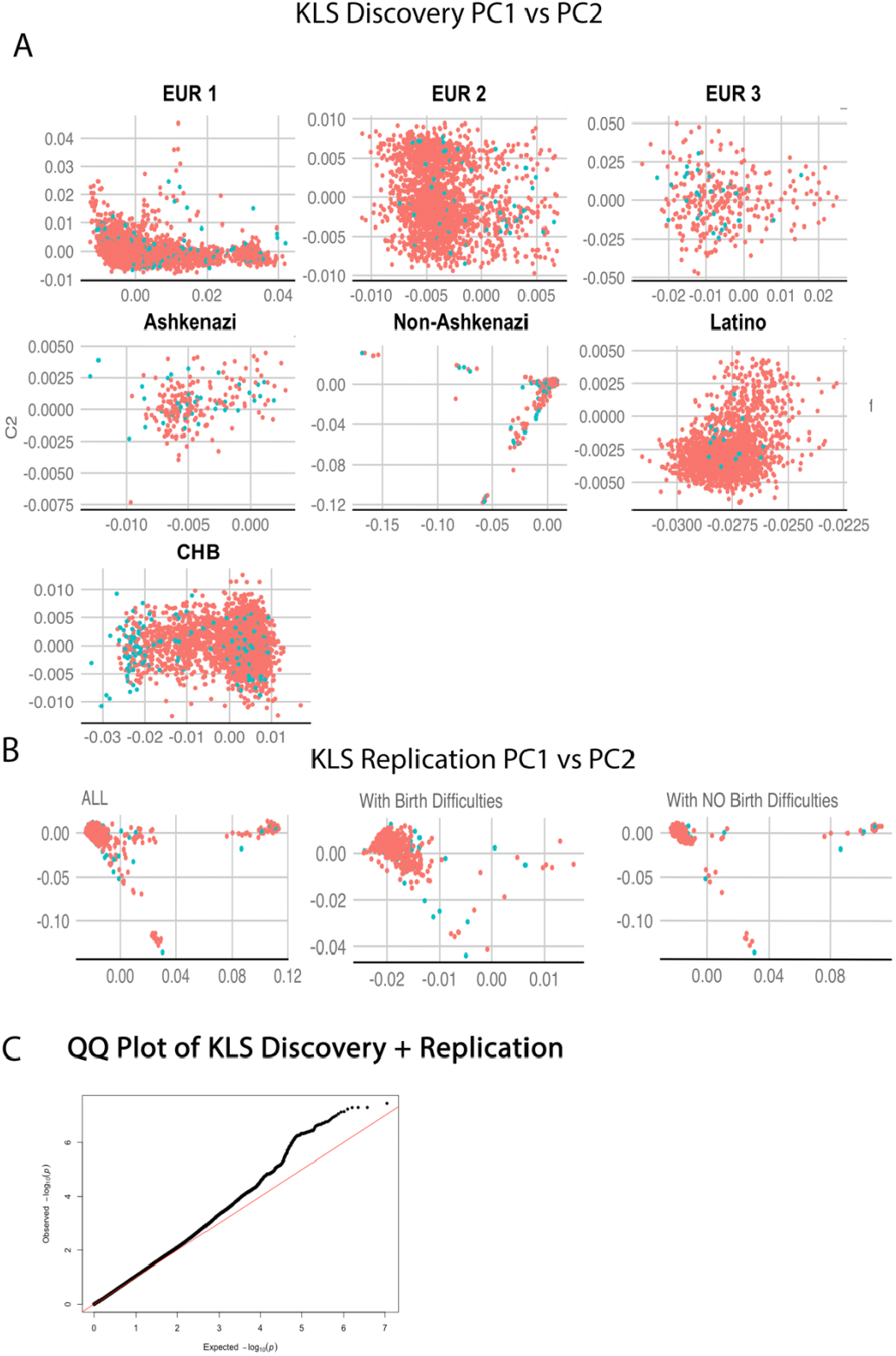
**A:** The first 2 genetic principle components matching of case-controls in KLS discovery and **(B)** replication sub -cohorts stratified by presence of birth difficulties. **(C)** QQplot of the total KLS case-controls combined discovery + replication genetic association p-values.

**Fig. S2.**
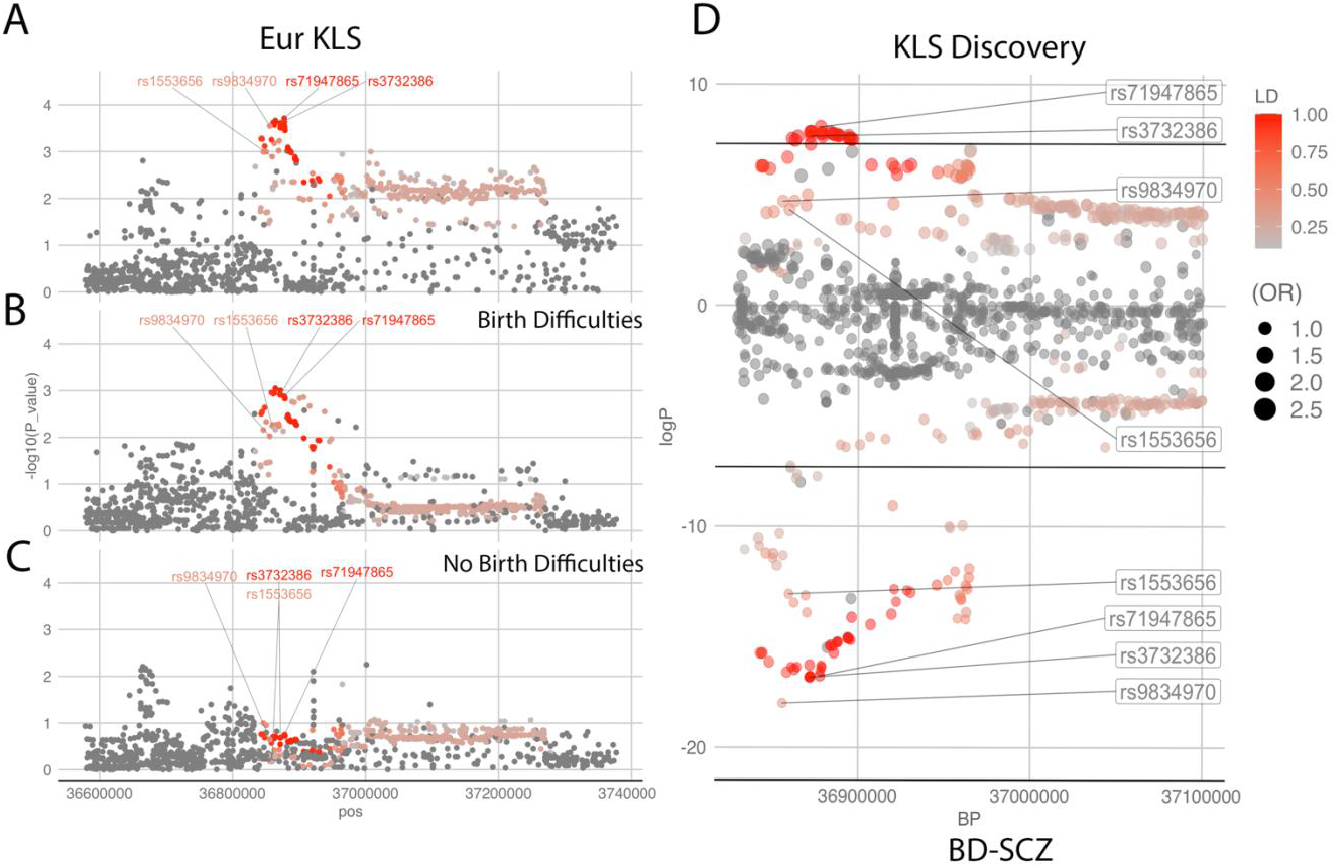
**(A-C):** TRANK1 regional plots in the European origin KLS meta-analysis and sub-cohorts with and with our birth difficulties. **(D)** – TRANK1 loci plots with KLS discovery in the positive fraction contrasted with Bipolar Disorder-Schizophrenia (BD-SCZ) GWAS summary stats in the negative fraction(1) (Ruderfer DM et al. 2018), horizontal lines indicate GWAS significance threshold (p=5e-8).

**Fig. S3.**
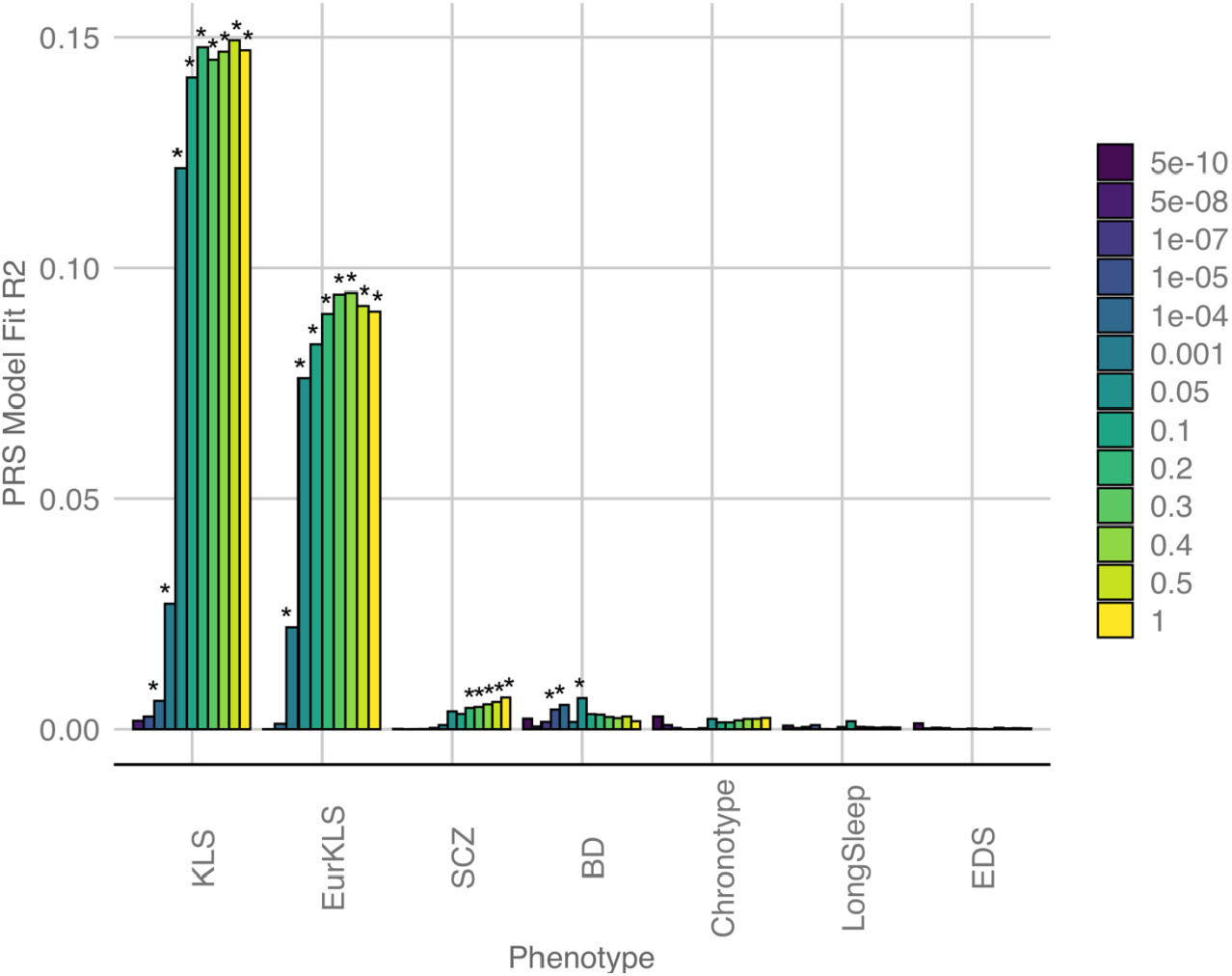
PRS (polygenic risk scores) constructed from various other GWAS studies i.e. bipolar disorder, schizophrenia, circadian chronotype, sleep duration and excessive daytime sleepiness, evaluating their predictive performance on the KLS follow up cohort. EurKLS – PRS built on Caucasian discovery and validated in Caucasian replication KLS samples. The y-axis is the % variance explained by the PRS while the x-axis represents the p-value thresholds. Inset are marked the Pvalue of the fit. The * on the top of the bars represent the empirical p value (p<0.05) associated with the test of p-value threshold.

**Dataset S1**: The total cohort used in the analysis split by sub-cohort and genotyping array used grouped by KLS cases and controls.

**Dataset S2**: Variants in the TRANK1 loci associated with KLS in worldwide discovery cohort GWAS meta-analysis in 673 KLS cases and 15341 control individuals and in a Follow-up replication cohort of 171 KLS cases and 1956 controls. The effect allele was B*.

**Dataset S3**: TRANK1 region SNPs correlation in schizophrenia and bi-polar disorder GWAS studies. * indicates Genome wide significant (p<5e-8). PGC is Psychiatric Genomics Consortium.

**Dataset S4**: TRANK1 deletion (rs71947865) and its univariate association with KLS clinical variables. We used 2 linear additive models Model1-the additively coded minor allele genotype was fit as a function of the clinical variable, Model2 – was similar to model1 except for including ethnicity, Gender and missing data proportion. A Fisher’s exact test of allelic counts was further performed for categorical variables. In these analyses 592 KLS cases were included. (Frequency % | mean ± sem was computed). BB-allele G, AB-GGGAGCCA/G, AA - GGGAGCCA/ GGGAGCCA, *Minor Allele G (deletion) was the effect allele.

**Dataset S5**: Birth Difficulty and developmental delay summary in the KLS cases.

**Dataset S6: (A)** TRANK1 SNPs in birth year stratified analysis, Fisher’s exact count test was performed for case -control analyses. Minor *A allele was effect allele. **(B)** Allelic counts of TRANK1 SNPs in KLS cases compared to each birth year bin, Fisher’s exact count test was performed for case only analyses. Minor A allele was effect allele.

**Dataset S7:** Variants in the TRANK1 loci associated with KLS in worldwide discovery cohort GWAS meta-analysis in 673 KLS cases and 15339 control individuals and Follow-up replication sub-cohorts based on presence of birth difficulties. The effect allele was B*. genomic coordinates in hg19/GRCh37.

**Dataset S8**: **(A)** TRANK1 SNPs association in KLS cases with birth difficulties compared to controls stratified by discovery and the replication cohort, Fisher’s exact count test was performed. Minor *A allele was effect allele. **(B)** TRANK1 SNPs in birth difficulty stratified analysis in the combined discovery and replication cohort, Fisher’s exact count test was performed. Minor *A allele was effect allele.

**Dataset S9**: Meta-analysis of Variants in the TRANK1 loci in European origin KLS cases in 2 cohorts stratified by all, presence of birth difficulties and absence of birth difficulties. Effect allele is B*. Filter on Analysis column to see associations by birth difficulties. genomic coordinates in hg19/GRCh37.

**Dataset S10**: Polygenic risk score analysis of KLS follow up sample in relation to other disorders including Bipolar Disorder, Schizophrenia, Circadian chronotype and Excessive daytime sleepiness.

**Dataset S11:** LD hub output containing LD score regression and shared SNP heritability with KLS across phenotypes, the table lists the top traits that share heritability with KLS.

**Dataset S12:** Top 2 enriched pathways in a MAGMA based gene set enrichment analysis was carried out using A total of 5346 sets including curated gene sets, KEGG pathways, Reactome pathways and GO biological processes derived from msigdb6.1 were used. Genomic coordinates in hg19/GRCh37.

**Dataset S13: (A)** The KLS cases and families (n=32) used in the exome sequencing studies. Controls were derived from 1000 genomes phase 3. **(B)**: Frequency and burden of all LMOD3 gene variants derived from whole exome sequencing in 17 KLS Probands/sporadic cases and 286 European controls. Genomic coordinates in hg19/GRCh37.

**Dataset S14: (A)** Exome burden test restricted to rare variants with high and moderate effects grouped by reference genes within each families, probands and total cohort are listed, Columns with prefix KLS implies all the 32 cases and column prefix proband includes the proband in each family in addition to the 3 sporadic cases (n=17). genomic coordinates in hg19/GRCh37.**(B):** Exome burden test restricted to rare variants with high and moderate effects grouped by reference genes shared by at least 3 families, Columns with prefix KLS implies all the 32 cases and column prefix proband includes the proband in each family in addition to the 3 sporadic cases (n=17). genomic coordinates in hg19/GRCh37.

**Dataset S15: (A)** Top hits from Exome SKAT model with population covariates in 32 KLS cases and 286 controls with a focus on collapsing variants in the genes belonging to REACTOME_RORA_ACTIVATES_CIRCADIAN_EXPRESSION and REACTOME_CIRCADIAN_REPRESSION_OF_EXPRESSION_BY_REV_ERBA both of which were 5% FDR significant in the joint Discovery-Replication KLS GWAS pathway analyses. **(B)**: Top hits from Exome SKAT model with population covariates in 17 KLS probands and 286 controls with a focus on collapsing variants in the genes in pathways derived from a total of 5346 sets including curated gene sets, KEGG pathways, Reactome pathways and GO biological processes derived from msigdb6.1 were used.

